# LightFusionNet: Lightweight Dual-Stream Network with Predictive Context Attention for Efficient Medical Image Fusion

**DOI:** 10.1101/2025.10.25.25338800

**Authors:** Abhinav Sagar

## Abstract

Multimodal image fusion aims to integrate complementary information from multiple imaging modalities into a single, informative representation, which is crucial for applications in medical imaging and microscopy. Existing methods often face trade-offs between structural fidelity, edge preservation, and computational efficiency. In this work, we propose LightFusionNet, a lightweight dual-stream network designed to efficiently fuse multimodal images while retaining key structural, textural, and intensity features. The network leverages depthwise separable convolutions to reduce model complexity and incorporates a Predictive Context Attention (PCA) mechanism to selectively emphasize informative regions in the feature maps. Extensive experiments on benchmark medical imaging datasets, including PET-MRI, SPECT-MRI, and CT-MRI, demonstrate that our approach achieves comparable qualitative and quantitative performance compared to state-of-the-art fusion methods, while maintaining low computational cost. The proposed method provides an effective and efficient solution for multimodal image fusion, suitable for both clinical and research applications.

## Introduction

Multimodal image fusion aims to integrate complementary information from multiple imaging modalities into a single, informative representation. In medical imaging, for instance, combining functional information from Positron Emission Tomography (PET) or Single-Photon Emission Computed Tomography (SPECT) with anatomical details from Magnetic Resonance Imaging (MRI) can enhance diagnostic accuracy and assist clinical decision-making.

Traditional image fusion methods, such as those based on multi-scale transforms or sparse representations, rely heavily on handcrafted features and manually designed fusion rules. While these approaches are computationally efficient, they often struggle to capture complex nonlinear relationships and fail to generalize well across diverse datasets. The emergence of deep learning has revolutionized image fusion by enabling end-to-end learning of feature extraction, fusion, and reconstruction. Autoencoder-based, CNN-based, and transformer-based architectures have demonstrated remarkable performance, effectively preserving salient structures, textures, and cross-modal information.

Despite these advancements, existing methods face several challenges. Autoencoder-based networks often rely on predefined fusion rules, limiting adaptability. CNN-based approaches, while effective at capturing local features, have restricted receptive fields, making it difficult to model long-range dependencies. Transformer-based models address global context modeling but typically incur high computational costs, which can hinder deployment in resource-constrained environments. Moreover, many existing networks are not optimized for balancing structural fidelity, edge preservation, and computational efficiency simultaneously.

In this work, we present an image fusion framework that achieves an optimal balance between fusion quality and computational efficiency. As illustrated in Figure 1, our model attains higher mutual information compared to existing methods while maintaining a significantly lower runtime and parameter count. The scatter visualization highlights that, unlike several high-performing but computationally heavy models, our approach delivers strong information preservation with efficient inference, making it well-suited for real-time and resource-constrained clinical applications.

**Figure 1:**
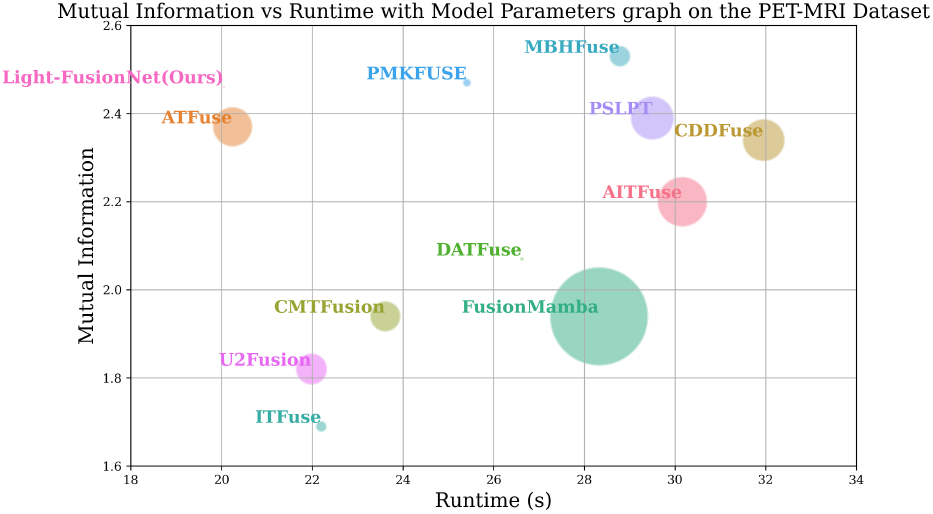
Comparison of various image fusion models in terms of mutual information on the Y axis, runtime (test set in seconds) on the X axis, and the number of parameters (in millions) represented by the area of the circle on the PET-MRI dataset. Our model achieves a good balance between performance and computational complexity.

The three main contributions of this paper are as follows:

1. We propose **LightFusionNet**, a lightweight dual-stream network for multimodal image fusion that efficiently integrates complementary features from each modality while preserving structural, textural, and intensity information.
2. We introduce a **Predictive Context Attention (PCA)** mechanism to selectively emphasize informative regions in the feature maps, enhancing the quality of fused images without increasing computational complexity.
3. Extensive experiments on benchmark medical imaging datasets, including PET-MRI, SPECT-MRI, and CT-MRI, demonstrate that our approach achieves comparable qualitative and quantitative performance compared to state-of-the-art methods, while maintaining low model complexity and high computational efficiency.

## Related Work

Recent progress in deep learning has significantly advanced image fusion, particularly in medical, infrared-visible, and multi-focus imaging. Existing methods can be broadly categorized into autoencoder-based, convolutional neural network (CNN)-based, and transformer-based paradigms. Each paradigm differs in its approach to feature extraction, representation learning, and fusion strategy, aiming to preserve complementary modality information while maintaining computational efficiency.

### Autoencoder-Based Image Fusion

Autoencoder-based approaches form the foundational paradigm for deep image fusion, employing encoder–decoder architectures to learn joint latent representations of multiple modalities. Fusion is typically achieved via pixel-level, feature-level, or decision-level integration in the latent space. DenseFuse (Li and Wu 2018) pioneered the use of DenseNet-style feature reuse to enhance representational richness and information flow between modalities. DeepFuse (Ram Prabhakar, Sai Srikar, and Venkatesh Babu 2017) introduced an unsupervised autoencoder for exposure fusion, using reconstruction loss to guide modality alignment. Similarly, U2Fusion (Xu et al. 2020) proposed a unified unsupervised architecture that preserves both global structure and modality-specific features through adaptive information preservation. These approaches are effective for general fusion tasks but rely heavily on predefined fusion rules, which can limit adaptability across modalities such as PET-MRI or infrared-visible pairs.

### CNN-Based Image Fusion

CNN-based models exploit hierarchical spatial feature extraction and local contextual modeling. Architectures like NestFuse (Li, Wu, and Durrani 2020) integrate dense connections and spatial–channel attention modules to enhance salient information, while IFCNN (Zhang et al. 2020) employs distinct feature extraction, fusion, and reconstruction blocks optimized via perceptual loss for improved texture and edge preservation. SDNet (Zhang and Ma 2021) introduces a squeeze-and-decomposition network for real-time image fusion, achieving a balance between performance and speed. Multi-task frameworks such as SuperFusion (Tang et al. 2022a) and Cross-UNet (Wang, Hua, and Li 2023) further enhance robustness across varied imaging domains.

Lightweight attention modules like SENet (Hu, Shen, and Sun 2018), CBAM (Woo et al. 2018), and ECA-Net (Wang et al. 2020) have also been widely adopted for adaptive feature recalibration, while Non-local networks (Wang et al. 2018) enable long-range dependency modeling within CNN backbones. Despite their success, CNNs remain limited in capturing long-distance contextual relationships crucial for multi-modal alignment.

### Transformer-Based Image Fusion

Transformers have emerged as a powerful alternative, addressing the limitations of CNNs by leveraging selfattention to capture global dependencies. The seminal Vision Transformer (ViT) (Dosovitskiy et al. 2021) and Swin Transformer (Liu et al. 2021) architectures inspired their adaptation to fusion tasks. In multimodal fusion, MATR (Tang et al. 2022c) and YDTR (Tang, He, and Liu 2022) employ multi-scale adaptive transformers for medical and infrared-visible fusion, respectively, improving feature correspondence between modalities. DATFuse (Tang et al. 2023b) and TCCFusion (Tang, He, and Liu 2023) utilize dual and cross-attention transformers to enhance complementary information exchange, while FATFusion (Tang and He 2024) integrates anatomical and functional cues for medical imaging. Similarly, Cross-Modal Transformer (Park, Vien, and Lee 2023) and CrossFuse (Li and Wu 2024) explore spatial–channel cross-attention to suppress redundant features and emphasize modality-specific details. Recent works like MixFuse (Li et al. 2025), AITFuse (Wang et al. 2024), and FusionMamba (Xie et al. 2024) employ hybrid transformer–state-space architectures for enhanced global context modeling and dynamic feature refinement. Moreover, CDDFuse (Zhao et al. 2023) introduces correlation-driven feature decomposition, and MBHFuse (Sun et al. 2025) incorporates differential convolution amplification to preserve structural consistency. These methods achieve strong performance but at the cost of higher computational complexity, motivating more lightweight yet expressive fusion networks.

### Hybrid and Advanced Architectures

Hybrid models combining CNN and transformer paradigms have gained traction for balancing local detail preservation with global context understanding. SwinFusion (Ma et al. 2022) integrates hierarchical transformer layers for longrange interaction, while PIAFusion (Tang et al. 2022b) introduces illumination-aware feature alignment. Recent developments like DCAFusion (Fang et al. 2025) and Rethinking Cross-Attention (Jian et al. 2024) further refine attention mechanisms to ensure consistent modality correspondence. State-space-inspired networks such as PMK-Fuse (Sun, Dong, and Zhu 2025) push toward lightweight global modeling by unifying KAN-based model locality and Mamba-based global context modeling.

## Discussion

Overall, these prior works highlight a fundamental trade-off between local feature preservation, global context modeling, and computational efficiency. CNN-based networks excel in local detail representation but struggle with long-range dependencies, while transformer-based models achieve superior global fusion at the expense of computational cost. The current research trend thus focuses on hybrid lightweight fusion frameworks that achieve an optimal balance among interpretability, accuracy, and efficiency—particularly important for medical image fusion tasks.

## Methodology

### Problem Definition

For PET-MRI and SPECT-MRI datasets, one modality is an RGB image (*I*_1_ ∈ *R*^*H*×*W*×3^), while the other is a grayscale image (*I*_2_ ∈ *R*^*H*×*W*×1^). The goal of fusion is to generate a single RGB fused image (*I*_*f*_ ∈ *R*^*H*×*W*×3^) that effectively integrates and preserves the essential information from both input modalities. To handle the channel inconsistency between the RGB and grayscale inputs, the RGB image is first converted to the YUV color space, separating it into Y (luminance), U (chrominance), and V (chrominance) components. The Y component is then combined with the grayscale image and passed through the fusion network. The final fused output is obtained by converting the result from YUV back to RGB.

For the CT-MRI dataset, both input images are grayscale (*I*_1_, *I*_2_ ∈ *R*^*H*×*W*×1^). The fusion objective here is to produce a single grayscale fused image (*I*_*f*_ ∈ *R*^*H*×*W*×1^) that captures complementary details from both modalities.

### Network Architecture

We propose **LightFusionNet**, a lightweight dual-stream architecture specifically designed for efficient medical image fusion. The network integrates modality-specific encoders, a *Predictive Context Attention (PCA)* fusion block, and a compact decoder to balance fusion quality with computational efficiency. By leveraging context prediction and attention-based refinement, the proposed design effectively preserves salient anatomical and functional information while minimizing redundant computations.

#### Encoder

Each modality—such as PET and MRI—is processed through an independent encoder constructed using *Depthwise Separable Convolutions (DSC)* (Chollet 2017). This decomposition of standard convolutions into depthwise and pointwise operations drastically reduces the number of parameters and floating-point operations (FLOPS) without compromising representation power. Each encoder progressively extracts hierarchical features using convolutional blocks with increasing receptive fields and a base channel width of 16. The resulting representations capture modality-specific textures and structural cues essential for downstream fusion.

#### Predictive Context Attention (PCA)

The PCA module serves as the key innovation of LightFusionNet, enabling dynamic context reasoning. For each modality, local spatial context is used to predict the expected feature response, and a *surprise map* is computed as the absolute deviation between predicted and actual features. This map identifies regions where contextual predictions fail—often corresponding to salient anatomical or functional transitions. A sigmoid activation transforms the surprise map into adaptive attention weights, amplifying informative or uncertain regions while suppressing redundant background responses. To ensure stability and gradient flow, a residual connection adds the weighted features back to the original input, forming a predictive feedback loop that enhances interpretability and robustness.

#### Fusion Block

The modality-specific attention-refined features are combined using an element-wise averaging operation, producing a compact joint representation. This simple yet effective strategy avoids introducing additional parameters while retaining complementary modality-specific cues. The fused representation is thus information-rich yet computationally lightweight, making it particularly suitable for real-time and embedded medical imaging applications.

#### Decoder

The decoder reconstructs the fused image from the integrated representation using a sequence of depthwise separable convolutions, followed by a 1 × 1 convolution and sigmoid activation.

Figure 2 summarizes the overall design of the proposed PET–MRI fusion framework. As shown in part A, the network adopts an encoder–decoder paradigm augmented with cross-modal predictive attention to integrate structural and functional information. Part B visualizes the PCA module’s adaptive context prediction and weighting mechanism, while part C highlights the efficiency of the DSC block. Collectively, these components enable LightFusionNet to achieve a favorable balance between fusion quality, interpretability, and computational economy, outperforming conventional heavy-weight architectures in both accuracy and runtime.

**Figure 2:**
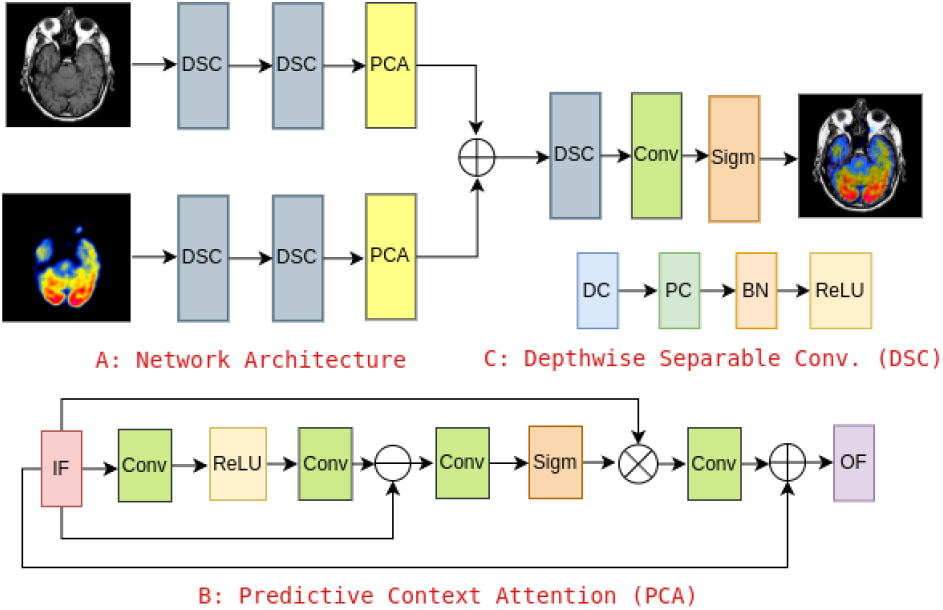
A: Overview of the proposed network architecture. B: The Predictive Context Attention (PCA) module predicts spatially adaptive attention weights from contextual deviations. C: Depthwise Separable Convolution (DSC) structure for efficient feature extraction.

### Loss Function

Our model is trained in an unsupervised manner using a combination of complementary loss functions that enforce intensity consistency, edge preservation, and structural fidelity in the fused images.

#### Intensity Loss

To ensure that the fused image retains the dominant pixel intensity information from the source images, we define an intensity loss as:

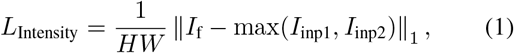

where *I*_f_ is the fused image, *I*_inp1_ and *I*_inp2_ are the source images, *H* and *W* denote the image height and width, and max is applied element-wise.

#### Gradient Loss

To preserve edge information and fine textures, we introduce a gradient-based loss computed using the Sobel operator:

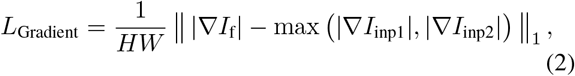

where ∇ denotes the gradient operator, and |·| represents the gradient magnitude.

#### Structural Similarity (SSIM) Loss

To retain structural information from the source images, we employ SSIM loss, which measures similarity in terms of luminance, contrast, and structure:

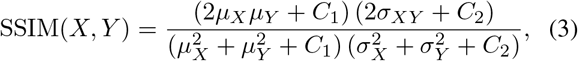

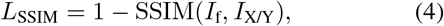

where *µ* and *σ* denote the mean and standard deviation, *σ*_*XY*_ is the covariance, and constants *C*_1_ = 0.01, *C*_2_ = 0.03 stabilize the calculation.

#### Total Loss

The overall loss is a weighted sum of the three components:

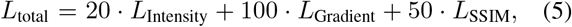

where the weights are empirically set to balance intensity, edge, and structural preservation during training.

## Experiments

### Dataset

To evaluate the performance of our proposed method, we conduct experiments on the following datasets:

1. **PET-MRI:** This dataset consists of 311 PET-MRI image pairs, with 269 images allocated for training and 42 images reserved for testing. All images have a resolution of 256 × 256 (Summers 2003).
2. **SPECT-MRI:** Comprising 430 SPECT-MRI image pairs, this dataset is divided into 357 training images and 73 testing images, each with a resolution of 256 × 256 (Summers 2003).
3. **CT-MRI:** This dataset contains 205 CT-MRI image pairs, split into 184 images for training and 21 images for testing. The image resolution is 256 × 256 (Summers 2003).

### Implementation Details

Our model is trained using the Adam optimizer with an initial learning rate of 10^−4^. The learning rate is adjusted using a MultiStepLR scheduler, which reduces it by a factor of 0.5 every 50 epochs. Training is performed with a batch size of 4 for a total of 200 epochs on an Nvidia A100 GPU. During training, the images are randomly cropped into patches of size 64 × 64 to facilitate data augmentation and improve convergence.

### Evaluation Metrics

To comprehensively evaluate the performance of our model, we adopt multiple metrics that capture different aspects of image fusion quality. **Entropy (EN)** quantifies the amount of information contained in the fused image, while **Standard Deviation (SD)** measures intensity variation. **Spatial Frequency (SF)** reflects edge activity, and **Average Gradient (AG)** assesses edge sharpness. Reference-based metrics include **Mutual Information (MI)**, which evaluates how much information from the source images is preserved; **Structural Content Difference (SCD)**, which measures structural consistency; and **Correlation Coefficient (CC)**, which indicates linear correlation between the fused and source images. For all metrics, higher values indicate better performance. Note that EN, SD, SF, and AG are no-reference metrics that do not require ground-truth images, whereas MI, SCD, and CC are reference-based metrics.

Computational efficiency is quantified using the number of model parameters (Param, in millions), floating point operations per second (FLOPS, in GigaFLOPS), and inference time (in seconds) on the test set, all computed on images of resolution 256 × 256. Lower values are preferred for Param, FLOPS, and inference time.

### Comparison Approaches

We compare our proposed model, with several state-of-the-art image fusion methods, including AITFuse (Wang et al. 2024), ATFuse (Jian et al. 2024), CDDFuse (Zhao et al. 2023), CMTFusion (Park, Vien, and Lee 2023), DATFuse (Tang et al. 2023b), FusionMamba (Xie et al. 2024), ITFuse (Tang, He, and Liu 2024), MBHFuse (Sun et al. 2025), PMKFuse (Sun, Dong, and Zhu 2025), PSLPT (Wang, Deng, and Vivone 2024), and U2Fusion (Xu et al. 2020). Performance results are reproduced using publicly available implementations provided by the authors, adhering to the experimental settings described in their respective papers. For PSLPT (Wang, Deng, and Vivone 2024), we adopt an unsupervised training strategy to ensure consistency with both our method and other comparative approaches.

#### Quantitative Performance

Table 1 summarizes the performance of our method in comparison with state-of-the-art fusion approaches across the PET-MRI, SPECT-MRI, and CT-MRI datasets. Our method consistently achieves comparable scores across multiple metrics, demonstrating that the fused images contain rich information and well-preserved edge details at a fraction of the computational cost of even the most lightweight model.

**Table 1:** A: Quantitative comparison on the PET-MRI dataset. B: Quantitative comparison on the SPECT-MRI dataset. C: Quantitative comparison on the CT-MRI dataset. The best values are highlighted in bold.

| A: PET-MRI Dataset |  |  |  |  |  |  |  |  |  |  |
| --- | --- | --- | --- | --- | --- | --- | --- | --- | --- | --- |
| Method | EN ↑ | SD ↑ | SF ↑ | AG ↑ | MI ↑ | SCD ↑ | CC ↑ | Param ↓ | FLOPS ↓ | Time ↓ |
| AITFuse (Wang et al. 2024) | **6.41** | 55.78 | 8.41 | **3.48** | 2.20 | 1.24 | 0.62 | 6.500 | 82.09 | 30.16 |
| ATFuse (Jian et al. 2024) | 4.01 | 71.41 | 8.22 | 2.86 | 2.37 | 1.22 | 0.61 | 1.050 | 5.40 | 20.24 |
| CDDFuse (Zhao et al. 2023) | 4.07 | 61.01 | 8.10 | 2.77 | 2.34 | **1.29** | **0.65** | 1.188 | 77.68 | 31.95 |
| CMTFusion (Park, Vien, and Lee 2023) | 4.04 | 47.64 | 6.73 | 2.40 | 1.94 | 1.19 | 0.59 | 0.624 | 13.10 | 23.61 |
| DATFuse (Tang et al. 2023b) | 4.18 | **87.38** | 7.45 | 2.53 | 2.07 | 1.19 | 0.59 | 0.010 | 2.32 | 26.62 |
| FusionMamba (Xie et al. 2024) | 4.30 | 67.80 | **8.73** | 3.06 | 1.94 | 1.20 | 0.60 | 225.420 | 26.48 | 28.32 |
| ITFuse (Tang, He, and Liu 2024) | 4.34 | 38.53 | 5.82 | 1.97 | 1.69 | 1.23 | 0.61 | 0.082 | 5.68 | 22.20 |
| MBHFuse (Sun et al. 2025) | 3.93 | 60.26 | 8.11 | 2.78 | **2.53** | 1.29 | 0.64 | 0.299 | 28.92 | 28.78 |
| PMKFuse (Sun, Dong, and Zhu 2025) | 3.91 | 61.42 | 8.10 | 2.75 | 2.47 | 1.28 | 0.64 | 0.049 | 3.29 | 25.41 |
| PSLPT (Wang, Deng, and Vivone 2024) | 4.06 | 66.94 | 8.07 | 2.79 | 2.39 | 1.26 | 0.63 | 1.256 | 24.56 | 29.50 |
| U2Fusion (Xu et al. 2020) | 4.49 | 47.00 | 5.28 | 2.02 | 1.82 | 1.20 | 0.60 | 0.659 | 43.17 | 21.98 |
| Ours(LightFusionNet) | 4.06 | 59.22 | 7.72 | 2.65 | 2.46 | 1.27 | 0.63 | **0.004** | **0.29** | **20.05** |
| B: SPECT-MRI Dataset |  |  |  |  |  |  |  |  |  |  |
| Method | EN ↑ | SD ↑ | SF ↑ | AG ↑ | MI ↑ | SCD ↑ | CC ↑ | Param ↓ | FLOPS ↓ | Time ↓ |
| AITFuse (Wang et al. 2024) | 4.09 | **58.38** | 7.02 | 2.30 | 3.07 | 1.70 | 0.85 | 6.500 | 82.09 | 51.76 |
| ATFuse (Jian et al. 2024) | 3.77 | 57.57 | 7.18 | 2.35 | 2.74 | 1.71 | 0.85 | 1.050 | 5.40 | 35.56 |
| CDDFuse (Zhao et al. 2023) | 3.79 | 58.27 | 7.08 | 2.28 | **3.13** | 1.69 | 0.85 | 1.188 | 77.68 | 58.31 |
| CMTFusion (Park, Vien, and Lee 2023) | 3.75 | 43.60 | 5.13 | 1.74 | 2.36 | **1.74** | **0.87** | 0.624 | 13.10 | 43.15 |
| DATFuse (Tang et al. 2023b) | **5.09** | 49.80 | 7.11 | 2.57 | 2.33 | 1.71 | 0.86 | 0.010 | 2.32 | 40.22 |
| FusionMamba (Xie et al. 2024) | 3.87 | 57.21 | **8.05** | **2.63** | 2.32 | 1.67 | 0.84 | 225.420 | 26.48 | 48.80 |
| ITFuse (Tang, He, and Liu 2024) | 4.72 | 48.75 | 5.51 | 1.91 | 2.20 | 1.70 | 0.85 | 0.082 | 5.68 | 42.32 |
| MBHFuse (Sun et al. 2025) | 3.87 | 55.79 | 7.26 | 2.41 | 2.55 | 1.10 | 0.55 | 0.299 | 28.92 | 48.49 |
| PMKFuse (Sun, Dong, and Zhu 2025) | 3.75 | 40.20 | 6.91 | 2.26 | 2.33 | 1.31 | 0.66 | 0.049 | 3.29 | 45.58 |
| PSLPT (Wang, Deng, and Vivone 2024) | 3.77 | 57.95 | 6.96 | 2.24 | 2.91 | 1.71 | 0.86 | 1.256 | 24.56 | 52.09 |
| U2Fusion (Xu et al. 2020) | 3.90 | 45.30 | 4.01 | 1.37 | 2.22 | 1.74 | 0.87 | 0.659 | 43.17 | 38.51 |
| Ours(LightFusionNet) | 3.79 | 57.49 | 6.72 | 2.15 | 2.97 | 1.72 | 0.86 | **0.004** | **0.29** | **33.71** |
| C: CT-MRI Dataset |  |  |  |  |  |  |  |  |  |  |
| Method | EN ↑ | SD ↑ | SF ↑ | AG ↑ | MI ↑ | SCD ↑ | CC ↑ | Param ↓ | FLOPS ↓ | Time ↓ |
| AITFuse (Wang et al. 2024) | 4.30 | **87.38** | 7.41 | 2.59 | 2.26 | 1.62 | 0.81 | 6.500 | 82.09 | 14.73 |
| ATFuse (Jian et al. 2024) | 4.36 | 81.66 | 7.84 | 2.85 | 2.32 | 1.60 | 0.80 | 1.050 | 5.40 | 10.04 |
| CDDFuse (Zhao et al. 2023) | 4.68 | 79.27 | 8.10 | 3.03 | 2.32 | 1.61 | 0.80 | 1.188 | 77.68 | 15.49 |
| CMTFusion (Park, Vien, and Lee 2023) | 4.60 | 57.14 | 6.88 | 2.47 | 2.35 | **1.68** | **0.84** | 0.624 | 13.10 | 11.60 |
| DATFuse (Tang et al. 2023b) | 4.13 | 83.00 | 7.36 | 2.53 | 2.11 | 1.58 | 0.79 | 0.010 | 2.32 | 11.09 |
| FusionMamba (Xie et al. 2024) | 4.32 | 81.87 | **8.92** | **3.24** | 1.88 | 1.61 | 0.80 | 225.420 | 26.48 | 13.82 |
| ITFuse (Tang, He, and Liu 2024) | **5.24** | 25.86 | 6.85 | 2.63 | 1.60 | 1.33 | 0.66 | 0.082 | 5.68 | 11.39 |
| MBHFuse (Sun et al. 2025) | 4.48 | 79.81 | 7.98 | 2.95 | **2.41** | 1.59 | 0.79 | 0.299 | 28.92 | 14.12 |
| PMKFuse (Sun, Dong, and Zhu 2025) | 4.54 | 78.53 | 7.88 | 2.92 | 2.30 | 1.61 | 0.80 | 0.049 | 3.29 | 13.06 |
| PSLPT (Wang, Deng, and Vivone 2024) | 4.69 | 81.36 | 7.81 | 2.89 | 2.33 | 1.61 | 0.81 | 1.256 | 24.56 | 14.27 |
| U2Fusion (Xu et al. 2020) | 4.89 | 44.85 | 4.50 | 1.67 | 1.99 | 1.63 | 0.81 | 0.659 | 43.17 | 10.40 |
| Ours(LightFusionNet) | 4.77 | 78.26 | 7.47 | 2.78 | 2.35 | 1.63 | 0.81 | **0.004** | **0.29** | **9.43** |

**Table 2:** A: Quantitative comparison on the PET-MRI dataset using a model trained on the SPECT-MRI dataset. B: Quantitative comparison on the SPECT-MRI dataset using a model trained on the PET-MRI dataset. The best values are highlighted in bold.

| A: PET-MRI Dataset |  |  |  |  |  |  |  |  |  |  |
| --- | --- | --- | --- | --- | --- | --- | --- | --- | --- | --- |
| Method | EN ↑ | SD ↑ | SF ↑ | AG ↑ | MI ↑ | SCD ↑ | CC ↑ | Param ↓ | FLOPS ↓ | Time ↓ |
| AITFuse (Wang et al. 2024) | 4.25 | 60.93 | 8.02 | 2.74 | 2.38 | **1.30** | **0.65** | 6.50 | 82.09 | 29.92 |
| ATFuse (Jian et al. 2024) | 4.02 | 73.21 | 8.12 | 2.80 | 2.20 | 1.19 | 0.59 | 1.05 | 5.40 | 19.13 |
| CDDFuse (Zhao et al. 2023) | 3.70 | 66.80 | 6.99 | 2.22 | 2.13 | 1.08 | 0.54 | 1.19 | 77.68 | 32.69 |
| CMTFusion (Park, Vien, and Lee 2023) | 4.06 | 49.32 | 6.39 | 2.42 | 1.93 | 1.18 | 0.59 | 0.62 | 13.10 | 23.53 |
| DATFuse (Tang et al. 2023b) | **5.38** | 55.99 | 7.92 | **3.02** | 1.88 | 1.21 | 0.60 | 0.01 | 2.32 | 22.57 |
| FusionMamba (Xie et al. 2024) | 4.04 | 69.68 | **8.56** | 2.96 | 1.95 | 1.14 | 0.57 | 225.42 | 26.48 | 27.85 |
| ITFuse (Tang, He, and Liu 2024) | 5.06 | 51.85 | 6.41 | 2.36 | 1.84 | 1.19 | 0.59 | 0.08 | 5.68 | 23.22 |
| MBHFuse (Sun et al. 2025) | 4.21 | **75.30** | 8.32 | 2.91 | 1.98 | 0.66 | 0.33 | 0.30 | 28.92 | 28.70 |
| PMKFuse (Sun, Dong, and Zhu 2025) | 3.98 | 58.04 | 7.96 | 2.73 | 1.96 | 0.77 | 0.39 | 0.05 | 3.29 | 26.54 |
| PSLPT (Wang, Deng, and Vivone 2024) | 3.97 | 65.73 | 8.00 | 2.75 | **2.39** | 1.27 | 0.63 | 1.26 | 24.56 | 28.22 |
| U2Fusion (Xu et al. 2020) | 4.28 | 49.23 | 5.53 | 2.10 | 1.84 | 1.19 | 0.59 | 0.66 | 43.17 | 21.74 |
| Ours(LightFusionNet) | 4.13 | 62.14 | 7.87 | 2.73 | 2.13 | 1.28 | 0.64 | **0.004** | **0.29** | **18.99** |

| B: SPECT-MRI Dataset |  |  |  |  |  |  |  |  |  |  |
| --- | --- | --- | --- | --- | --- | --- | --- | --- | --- | --- |
| Method | EN ↑ | SD ↑ | SF ↑ | AG ↑ | MI ↑ | SCD ↑ | CC ↑ | Param ↓ | FLOPS ↓ | Time ↓ |
| AITFuse (Wang et al. 2024) | **5.66** | 55.72 | 7.32 | **2.83** | 2.62 | 1.67 | 0.83 | 6.50 | 82.09 | 51.72 |
| ATF (Jian et al. 2024) | 3.80 | 61.33 | 7.25 | 2.37 | 2.84 | 1.70 | 0.85 | 1.05 | 5.40 | 35.88 |
| CDDFuse (Zhao et al. 2023) | 3.84 | 60.10 | 7.02 | 2.27 | 2.84 | 1.72 | 0.86 | 1.19 | 77.68 | 58.10 |
| CMTFusion (Park, Vien, and Lee 2023) | 3.66 | 41.59 | 5.58 | 1.75 | 2.32 | **1.74** | **0.87** | 0.62 | 13.10 | 42.37 |
| DATFuse (Tang et al. 2023b) | 4.10 | **75.90** | 6.84 | 2.24 | 2.45 | 1.72 | 0.86 | 0.01 | 2.32 | 41.45 |
| FusionMamba (Xie et al. 2024) | 4.07 | 59.75 | **8.17** | 2.70 | 2.30 | 1.69 | 0.84 | 225.42 | 26.48 | 52.91 |
| ITFuse (Tang, He, and Liu 2024) | 3.91 | 36.73 | 4.96 | 1.46 | 2.01 | 1.71 | 0.85 | 0.08 | 5.68 | 42.10 |
| MBHFuse (Sun et al. 2025) | 3.71 | 56.60 | 7.10 | 2.30 | **3.11** | 1.70 | 0.85 | 0.30 | 28.92 | 52.64 |
| PMKFuse (Sun, Dong, and Zhu 2025) | 3.81 | 63.35 | 7.16 | 2.32 | 2.90 | 1.70 | 0.85 | 0.05 | 3.29 | 47.62 |
| PSLPT (Wang, Deng, and Vivone 2024) | 3.83 | 57.90 | 7.03 | 2.27 | 2.83 | 1.71 | 0.85 | 1.26 | 24.56 | 50.95 |
| U2Fusion (Xu et al. 2020) | 4.06 | 43.77 | 3.72 | 1.27 | 2.24 | 1.74 | 0.86 | 0.66 | 43.17 | 37.13 |
| Ours(LightFusionNet) | 3.73 | 53.85 | 6.70 | 2.13 | 2.63 | 1.71 | 0.85 | **0.004** | **0.29** | **36.88** |

#### Qualitative Performance

We evaluate the qualitative performance of our proposed method by visually comparing the fused images against the source images and the outputs of state-of-the-art fusion methods. The assessment focuses on three key aspects: preservation of salient features, retention of edge details, and maintenance of structural and intensity information.

Figures 3, 4, and 5 illustrate representative results on the PET-MRI, SPECT-MRI, and CT-MRI datasets. Our method consistently produces fused images that retain the informative regions from both source modalities. Specifically, it preserves high-intensity structures from the one image while simultaneously maintaining the fine texture and contrast from the other image.

**Figure 3:**
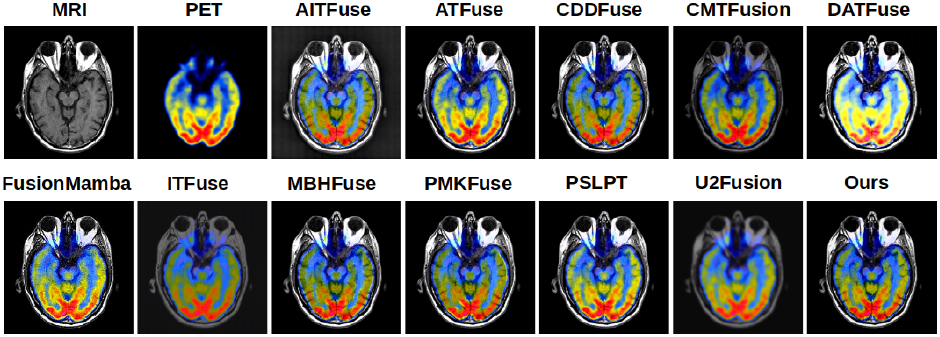
Qualitative comparison on the PET-MRI dataset with other state-of-the-art methods.

**Figure 4:**
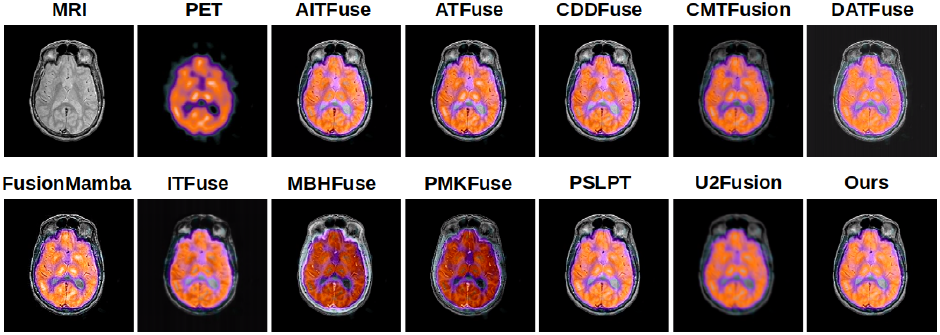
Qualitative comparison on the SPECT-MRI dataset with other state-of-the-art methods.

**Figure 5:**
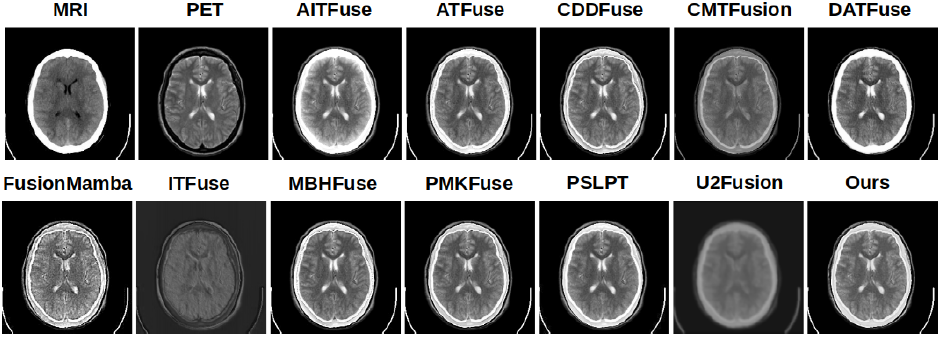
Qualitative comparison on the CT-MRI dataset with other state-of-the-art methods.

Compared to other approaches, the proposed method demonstrates superior clarity in regions with high-frequency details, such as edges and organ boundaries, while avoiding common artifacts such as blurring or over-saturation. The predictive context attention mechanism enables selective enhancement of informative regions, resulting in fused images that are both visually appealing and faithful to the original sources.

The ability of an image fusion model to generalize to unseen datasets without fine-tuning is crucial for real-world applications. However, most existing fusion approaches are tailored to specific datasets and tend to perform poorly when evaluated on out-of-distribution data. To assess the generalization capability of our method, we conduct cross-dataset experiments: the model trained on the SPECT-MRI dataset is tested on the PET-MRI dataset, and vice versa. The quantitative results of these evaluations are presented in 2. While our method does not consistently achieve the top performance across all metrics, it consistently ranks within the top three on multiple evaluation criteria, demonstrating strong generalization ability.

The qualitative results for cross-dataset evaluation are presented in Figure 6 and Figure 7, where the model trained on the SPECT-MRI dataset is tested on the PET-MRI dataset and vice versa. A noticeable decline in performance is observed for several existing methods, including CDDFuse, MBHFuse, and PMKFuse, which otherwise perform well when trained and tested on the same dataset. In contrast, our method exhibits stronger generalization capability, maintaining superior visual quality compared to most competing approaches at a fraction of the computational cost.

**Figure 6:**
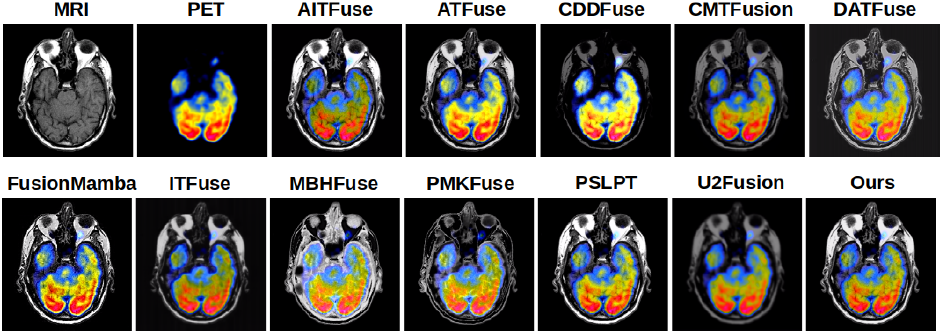
Qualitative comparison on the PET-MRI dataset using a trained model from the SPECT-MRI dataset with other state-of-the-art methods.

**Figure 7:**
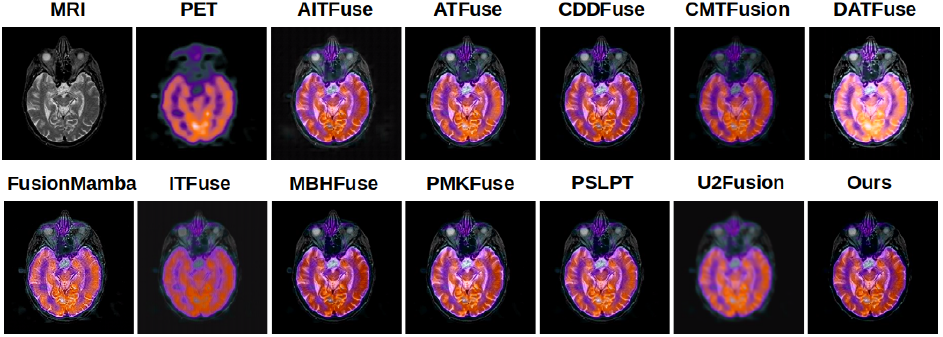
Qualitative comparison on the SPECT-MRI dataset using a trained model from the PET-MRI dataset with other state-of-the-art methods.

#### Ablation Study

To investigate the contributions of key components in **LightFusionNet**, we conduct an ablation study on the PET-MRI dataset. Specifically, we evaluate the impact of different attention mechanisms in the Predictive Context Attention (PCA) module and the use of depthwise separable (D/W) convolutions on overall fusion performance, computational cost, and inference time.

Table 3 summarizes the quantitative results. Replacing PCA with standard convolution (PCA → Conv) results in a noticeable drop in entropy (EN) and standard deviation (SD), indicating less informative and less contrast-rich fused images. Using Channel Attention (CA) in place of PCA improves SD and spatial frequency (SF), suggesting better preservation of structural details, while Spatial Attention (SA) performs slightly worse than CA.

**Table 3:** Ablation study using different components in the network architecture. Here Conv. denotes regular convolutional layer, CA denotes Channel Attention, SA denotes Spatial Attention, and D/W denotes depthwise convolution. The best values are highlighted in bold.

| Network | EN ↑ | SD ↑ | SF ↑ | AG ↑ | MI ↑ | SCD ↑ | CC ↑ | Param ↓ | FLOPS ↓ | Time ↓ |
| --- | --- | --- | --- | --- | --- | --- | --- | --- | --- | --- |
| PCA → Conv | 3.99 | 61.35 | 7.70 | 2.64 | 2.12 | 1.27 | 0.63 | **0.002** | 0.14 | **19.31** |
| PCA → CA | 4.04 | **65.91** | **7.97** | 2.59 | 2.16 | 1.27 | 0.62 | 0.002 | **0.11** | 20.41 |
| PCA → SA | 4.00 | 59.35 | 7.73 | 2.56 | 2.16 | 1.26 | 0.63 | 0.002 | 0.12 | 20.39 |
| D/W → Conv | 4.01 | 58.10 | 7.89 | 2.62 | 2.15 | 1.27 | 0.63 | 0.010 | 0.66 | 19.63 |
| Ours | **4.06** | 59.22 | 7.72 | **2.65** | **2.16** | **1.27** | **0.63** | 0.004 | 0.29 | 20.05 |

Similarly, replacing depthwise separable convolutions with standard convolutions (D/W → Conv) increases the number of parameters and FLOPS significantly, while achieving only marginal improvements in some fusion metrics.

Our proposed configuration (**Ours**), combining PCA with depthwise separable convolutions, achieves a balanced performance across all metrics, maintaining competitive EN, SD, and SF values while keeping model parameters, FLOPS, and inference time low. These results demonstrate that both the PCA module and the use of lightweight depthwise convolutions are critical for achieving efficient and high-quality multimodal image fusion.

Table 4 presents the quantitative ablation study analyzing the impact of embedding channel size and attention reduction ratio on PET-MRI fusion performance. In part A, increasing the embedding channels from 8 to 16 improves multiple metrics, with the best results achieved at 16 channels. This setting balances representational capacity and computational efficiency, while further increasing to 32 channels offers marginal gains at a higher cost. In part B, varying the attention reduction ratio shows that R = 4 yields the optimal balance between feature refinement and model complexity, outperforming both smaller (R = 2) and larger (R = 8) ratios. Overall, the results indicate that moderate embedding dimensionality and attention compression provide the best trade-off between performance and efficiency for the proposed PET-MRI fusion framework.

**Table 4:** Quantitative ablation study on the PET-MRI dataset. A: Number of Embedding Channels. B: Reduction-Ratio in the attention module. The best-performing values are highlighted in bold.

| A: Embedding Channel |  |  |  |  |  |  |  |  |  |  |
| --- | --- | --- | --- | --- | --- | --- | --- | --- | --- | --- |
| C | EN ↑ | SD ↑ | SF ↑ | AG ↑ | MI ↑ | SCD ↑ | CC ↑ | Param ↓ | FLOPS ↓ | Time ↓ |
| 8 | 3.88 | 57.63 | 7.41 | 2.54 | 2.04 | 1.21 | **0.64** | **0.003** | **0.25** | **18.72** |
| 16 | 4.06 | **59.22** | **7.72** | 2.65 | **2.16** | **1.27** | 0.63 | 0.004 | 0.29 | 20.05 |
| 32 | **4.10** | 58.47 | 7.66 | **2.68** | 2.12 | 1.26 | 0.62 | 0.008 | 0.46 | 23.91 |

| B: Attention Reduction Ratio |  |  |  |  |  |  |  |  |  |  |
| --- | --- | --- | --- | --- | --- | --- | --- | --- | --- | --- |
| R | EN ↑ | SD ↑ | SF ↑ | AG ↑ | MI ↑ | SCD ↑ | CC ↑ | Param ↓ | FLOPS ↓ | Time ↓ |
| 2 | **4.10** | 58.39 | 7.61 | 2.60 | 2.11 | 1.25 | 0.61 | 0.005 | 0.33 | 21.47 |
| 4 | 4.06 | **59.22** | **7.72** | **2.65** | **2.16** | 1.27 | **0.63** | 0.004 | 0.29 | 20.05 |
| 8 | 3.94 | 57.20 | 7.33 | 2.51 | 2.00 | **1.29** | 0.59 | **0.003** | **0.24** | **18.38** |

Table 5 shows the effect of different SSIM, Gradient, and Intensity loss weight combinations on PET-MRI fusion performance. A balanced configuration of SSIM = 20, Gradient = 50, and Intensity = 100 achieves the best overall results. This setting provides an effective trade-off between structural preservation, edge detail, and intensity consistency, demonstrating that proportional weighting among the three losses is crucial for optimal fusion quality.

**Table 5:** Quantitative ablation study on the PET-MRI dataset, evaluating the impact of different loss function components: SSIM, Gradient, and Intensity. The best-performing values are highlighted in bold.

| SSIM | Gradient | Intensity | EN ↑ | SD ↑ | SF ↑ | AG ↑ | MI ↑ | SCD ↑ | CC ↑ |
| --- | --- | --- | --- | --- | --- | --- | --- | --- | --- |
| 10 | 50 | 100 | 3.92 | 57.10 | 7.36 | 2.52 | **2.19** | 1.20 | 0.60 |
| 30 | 50 | 100 | 3.98 | 58.03 | 7.48 | 2.58 | 2.09 | 1.23 | 0.61 |
| 20 | 30 | 100 | 3.94 | 57.68 | **7.80** | 2.56 | 2.06 | 1.22 | **0.63** |
| 20 | 70 | 100 | 3.99 | 58.14 | 7.55 | 2.59 | 2.10 | 1.24 | 0.62 |
| 20 | 50 | 50 | 3.90 | 56.82 | 7.30 | 2.50 | 1.98 | 1.18 | 0.59 |
| 20 | 50 | 200 | **4.04** | 58.43 | 7.66 | 2.61 | 2.13 | 1.26 | 0.62 |
| 20 | 50 | 100 | 4.01 | **59.22** | 7.72 | **2.65** | 2.16 | **1.27** | 0.61 |

## Conclusions

In this work, we propose **LightFusionNet**, a lightweight and efficient network for multimodal image fusion, capable of integrating complementary information from various medical imaging modalities. The network leverages depthwise separable convolutions for computational efficiency and employs a Predictive Context Attention mechanism to selectively emphasize informative regions, preserving structural details, edge information, and intensity features. Extensive experiments on multiple benchmark datasets, including PET-MRI, SPECT-MRI, and CT-MRI dataset, demonstrate that our method achieves comparable qualitative and quantitative performance compared to state-of-the-art approaches, while maintaining low model complexity. Future work will focus on extending the framework to incorporate additional modalities, exploring self-supervised and uncertainty-aware learning strategies, and further improving fusion quality in challenging scenarios such as low-contrast or noisy inputs.

## Data Availability

All data produced are available online at https://datasets.simula.no/kvasir/

https://datasets.simula.no/kvasir/

